# Investigating Pre-Operative Joint-Level Biomechanics in Partial versus Total Knee Arthroplasty

**DOI:** 10.1101/2025.09.21.25336261

**Authors:** Madeline Rao Shishis, Fatima Gafoor, Matthew C. Ruder, Vincenzo E Di Bacco, Kim Madden, Anthony Adili, Dylan Kobsar

## Abstract

**Objective:** The purpose of this study was to investigate preoperative joint-level kinematics across multiple functional tasks including preferred-pace walking, fast-paced walking, and sit-to-stand, alongside self-reported pain and function in cohorts scheduled for partial (PKA) versus total knee arthroplasty (TKA).

**Methods:** Patients with end-stage knee osteoarthritis were recruited from St. Joseph’s Healthcare Hamilton. Self-reported measures including Oxford Knee Score, self-reported average pain, quality of life, and depression were collected. Functional tasks including preferred-pace walking, fast-paced walking, and sit-to-stand were collected and recorded using markerless motion capture. Joint-level kinematics were evaluated using linear mixed models using main effects of surgery type and walking condition, along with interaction effects.

**Results:** This study included 15 patients scheduled for PKA, and 56 patients scheduled for TKA. No significant differences were observed when evaluating differences between cohorts through self-reported measures, nor at single gait speed conditions or sit-to-stand. However, when evaluating the differences in walking patterns between preferred and fast pace, the PKA group exhibited a significant change in stride length, peak stance knee flexion, peak swing knee flexion, knee excursion, peak stance hip flexion, and overall hip range of motion between preferred and fast-paced walking compared to the TKA group.

**Conclusion:** These findings highlight that multi-speed gait assessments rather than single-speed walking or sit-to-stand tasks are likely a more sensitive approach for detecting biomechanical differences in OA patients and may have broader applicability to other clinical populations.

## Introduction

Osteoarthritis (OA) is the third most common chronic disease among older adults in Canada^1^. OA is commonly found in the knee joint, affecting approximately 37% of adults over the age of 60 years^2^. Patients with advanced knee OA walk with significant deficits in knee joint kinematics and kinetics^3^. As OA severity progresses, patients may experience a reduced gait speed, shorter stride length, and less range of motion (ROM) of the knee, hip, and ankle joints^4^. The economic burden of OA is substantial, costing the Canadian healthcare system approximately $1.4 billion (CAD) annually^5^. Over 75,000 knee replacement surgeries are performed annually in Canada, with projections of a 300 – 600% increase by 2030 in North America^5,6^. Given the high volume and projected increase in procedures, knee replacement surgery remains an important treatment option for individuals with end-stage knee OA.

Total Knee Arthroplasty (TKA) has been reported to show improvements in objective functional measures during preferred-pace walking between pre-operative and post-operative data collections^7^. Hatfield and colleagues reported an increased knee flexion angle during swing phase, decreased knee adduction moments during stance phase, and increased difference between the first peak and midstance knee adduction moment compared to pre-operative measurements^7^. However, it has also been reported that TKA patients experience lower peak knee flexion during stance and swing phase, lower ROM, lower external knee flexion moments, and lower external knee adduction moments when compared to healthy controls beyond one-year post-surgery^8^. Notably, only approximately 17% of patients undergoing TKA present with tricompartmental OA, underscoring the need for more personalized treatment options^9^.

Partial Knee Arthroplasty (PKA) is an increasingly common alternative to TKA, where one (unicompartmental – UKA) or two (bi-compartmental – BKA) compartments of the knee joint are replaced. This is a viable option if the OA-affected tissue is localized to one or two compartments of the knee. Studies have reported that PKA is a less invasive procedure with shorter operative times, faster post-operative rehabilitation, and fewer medical complications, which may ultimately translate to lower costs compared to TKA^10–12^. PKA has been reported to have statistically significant improvements in patient-reported outcomes over TKA early in recovery around 6-months post-surgery^13^, as well as late in recovery beyond 3-years post-surgery. However, other studies report no differences in patient-reported outcomes between PKA and TKA cohorts at 1-year post-surgery.

Although PKA candidates present with differing compartmental severity, self-reported pain and function preoperatively are remarkably similar to those of TKA candidates^13,14^. However, most studies rely exclusively on patient-reported outcomes and lack objective functional measures to quantify possible differences between PKA and TKA cohorts. There are some studies that evaluate objective functional measures between PKA and TKA cohorts through gait kinematics^15–17^, spatiotemporal parameters^16–20^, and clinical tests such as timed-up-and-go and 2-minute-walk-test^21^. Studies that report biomechanical differences showed that PKA may have improved ROM, side-to-side kinematic symmetry, knee extension during stance phase, greater gait speed and step time when compared to TKA at various time points post surgery^15,18,19,22,23^. Conversely, some studies report no differences between PKA and TKA regarding spatiotemporal parameters, joint angles, and joint moments, and some report larger varus in PKA knees^19,24^. However, very few of these studies include pre-operative data to compare these cohorts^16,20,21^. Furthermore, only one of these studies involves biomechanical tasks beyond a single-speed gait evaluation^19^. This highlights the need for more comprehensive methods to objectively assess preoperative functional performance in order to identify disparities between cohorts prior to surgery.

Based on the gaps in the literature, the purpose of this study was to investigate preoperative biomechanics across multiple functional tasks including preferred-pace walking, fast-paced walking, and sit-to-stand, alongside self-reported pain and function in cohorts that received TKA versus PKA. It was hypothesized that no differences would be observed between cohorts for self-reported measures, and patients that received PKA would show higher functionality during preoperative functional tasks compared to patients that received TKA.

## Materials and Methods

### Participants

This study is a secondary analysis of a prospective cohort study, which has been reviewed and approved by the Hamilton Integrated Research Ethics Board (HiREB 16236). Patients were recruited from the Fracture Clinic at St. Joseph’s Healthcare Hamilton from November 2023 to December 2024. Patients were informed about the study by the research staff during the patient’s preoperative assessment visit approximately one month prior to surgery (average 20 days ± 32 days). Informed consent was obtained prior to enrollment in the study. Patients were included in this study if they were aged 18 or older, diagnosed with end-stage OA, and received either a PKA or a TKA at St. Joseph’s Healthcare Hamilton. Patients were excluded from this study if they had significant disability preventing ambulation, were unable to provide informed consent, or could not complete all functional tasks.

While some patients were diagnosed with bilateral OA, surgical procedures were performed unilaterally. Surgical procedures were performed by two surgeons, one of whom conducted both PKA and TKA, while the other performed only TKA. The surgical decision to perform a PKA or a TKA was based on radiographic classification (x-ray), with final decisions left to the surgeon intraoperatively.

### Data Collection

Patient demographics including age, sex, and body mass index (BMI) were collected through a survey that was recorded and stored in the Research Electronic Data Capture (REDCap) database at McMaster University. Patient-reported outcome measures such as Oxford Knee Score (OKS), self-reported average pain (ratings from one week to time of data collection), quality of life (EQ-5D-5l), and depression (PHQ-8) were also collected through the REDCap database. The OKS is a 12-item questionnaire that evaluates self-reported function, with lower scores indicating lower function, and higher scores indicating higher function^25^. OKS data was missing for one question of one patient and was left blank. Pain questions such as “Rate your average pain in the last week” were evaluated using a visual analog scale from 0 (no pain) to 10 (worst pain imaginable). EQ-5D-5l assesses health across mobility, self-care, usual activities, pain/discomfort, and anxiety/depression, which is then transformed to an index value that represents how good (higher index) or bad (lower index) a person’s health state is^26^. EQ-5D-5l data was missing for one question for two patients and were not included in analysis. PHQ-8 is an 8-question evaluation of depression, with higher total scores representing more severe depression, and lower scores representing more minimal depression^27^. PHQ-8 data was missing for one question for two patients and were left blank. Patients also completed biomechanical functional tasks during their preoperative visit. Biomechanical functional tasks included 30-second quiet standing, 60-second preferred-pace walking, 30-second fast-paced walking, and 5 repetitions of sit-to-stand. For walking tasks, patients were instructed to walk along a 7.6-meter section of the hospital hallway. Video data were recorded using ten synchronized Sony Cyber-shot DSC-RX0 II digital cameras set up along the hospital hallway.

### Data Analysis

Video data was processed using markerless motion capture (V2023.1.0.3161, Theia Markerless, Kingston, ON, Canada). Biomechanical analysis was performed in Visual3D (C-Motion Inc., Kingston, ON) using a custom pipeline^28^. Walking data was segmented into individual strides, and spatiotemporal and kinematic variables were calculated for each stride. For sit-to-stand, the pelvis velocity was used to track the number of trials, with the actual test identified between the first and last stand in order to account for delayed attempts to sit on the last trial. Spatiotemporal and kinematic variables were calculated for each sit-to-stand trial.

Kinematic data and spatiotemporal parameters for each patient were extracted for further analysis in Python 3.12 using Visual Studio Code as the integrated development environment. A custom Python script was used to identify surgical side for each patient. Average sagittal knee, hip, and ankle angle waveforms during walking trials were plotted for PKA and TKA cohorts, time normalized to 100% gait. Computed differences between the fast-paced and preferred-pace waveforms were also plotted, where preferred-pace trials were subtracted from the fast-paced trials. Discrete measures were extracted, including gait speed (m/s), stride length (m), stride width (m), peak stance knee flexion (°), peak swing knee flexion (°), knee excursion (°), peak stance hip flexion (°), peak swing hip flexion (°), peak stance hip extension (°), overall hip ROM (°), overall peak ankle dorsiflexion (°), overall peak ankle plantarflexion (°), and overall ankle ROM (°). For sit-to-stand, mean time (s), peak knee flexion angles (°), minimum knee flexion angles (°), and peak trunk flexion angles (°) in the sagittal plane were extracted.

### Statistical Analysis

Statistical analysis was carried out in Python 3.12 using Visual Studio Code as the integrated development environment. For walking trials, linear mixed models were used to assess the main effects of surgery type (PKA or TKA), walk condition (preferred or fast), and the interaction between these two main effects, with patient ID set as the random effect. Standardized beta estimates were evaluated using z-scores, which allowed variables to be interpreted on the same scale using small (β = 0.2 – 0.5), medium (β = 0.5 – 0.8), and large (β > 0.8) effect sizes. For sit-to-stand, Student’s Independent Samples T-Tests (p < 0.05) were used to assess whether mean differences between PKA and TKA cohorts were statistically significant. Cohen’s d was used to evaluate small (d = 0.2 – 0.5), medium (d = 0.5 – 0.8), and large (d > 0.8) effect size. The Shapiro-Wilk Normality Test was used to test normal distribution for each discrete measure (p < 0.05 indicating non-normal distribution). When Normality Test failed (p < 0.05), the Mann-Whitney U Independent Samples T-Test was used to test statistical significance (p < 0.05), and rank biserial correlation was used to evaluate effect size.

## Results

104 patients were recruited for this study. 30 patients were excluded as they were unable to complete all biomechanical tasks, and a further 3 participants were removed due to unusable data during processing (camera drops, loose fitting clothing). A total of 71 patients were included in this study, with 15 patients that received PKA and 56 patients that received TKA. For patients who received a PKA, the medial and patellofemoral compartments were replaced in 11 patients, and the patellofemoral compartment was replaced in 4 patients.

For patients that were included in the study, comorbidities such as presence of other joint injury or surgery, and relevant medical conditions were collected (missing: n = 2). These comorbidities include previous traumatic knee injury (PKA: 1, TKA: 5), previous knee surgery (PKA: 2, TKA: 13), previous hip surgery (PKA: 0, TKA: 3), back pain (PKA: 6, TKA: 14), joint replacement of the upper body/back (PKA: 1, TKA: 0), OA in other joints (PKA: 7, TKA: 22), and rheumatoid arthritis (PKA: 1, TKA: 2).

Patient demographics (Table 1) revealed a statistically significant difference in age between PKA and TKA cohorts, where the PKA group was significantly younger (p < 0.001). Self-reported measures (Table 1), including OKS, pain, quality of life, and depression, showed no statistically significant differences between groups.

**Table 1.** Patient demographics and self-reported measures, with statistical significance (p<0.05) in bold. Values are reported as mean (standard deviation). Low OKS scores indicate lower function, and higher scores indicate higher function. Pain ratings were from 0 (none) to 10 (worst). EQ-5D-5l low index represents lower quality of life rating, and higher index represents higher quality of life rating. PHQ-8 indicate lower depression rating, and higher values indicate

|  | **PKA**<br>**(n = 15)** | **TKA**<br>**(n = 56)** | **p-value** |
| --- | --- | --- | --- |
| **Mean Age (years)** | **60 (7)** | **68 (9)** | **< .001** |
| Sex (% Female) | 73% Female | 55% Female | 0.21 |
| Mean BMI (kg/m<sup>2</sup>) | 32.5 (6.2) | 32.7 (6.7) | 0.92 |
| Oxford Knee Score | 25.7 (7.8) | 24.6 (7.7) | 0.64 |
| Average Pain in Last Week (/10) | 6.2 (2.0) | 6.3 (1.8) | 0.82 |
| Average Pain in Last 24 Hours (/10) | 6.1 (2.6) | 5.9 (2.0) | 0.80 |
| Average Worst Pain in 24 Hours (/10) | 7.5 (2.6) | 7.7 (2.1) | 0.78 |
| Average Pain at Time of Data Collection (/10) | 5.3 (3.0) | 4.2 (2.7) | 0.19 |
| Quality of Life (EQ-5D-5l) | 0.6 (0.2) | 0.7 (0.1) | 0.47 |
| Depression (PHQ-8) | 6.47 (4.8) | 4.71 (3.5) | 0.12 |

Waveforms for average knee, hip, and ankle flexion angles in the sagittal plane were plotted for PKA and TKA cohorts to compare preferred-pace walking and the difference between fast-paced and preferred-pace walking on the surgical side over 100% gait cycle (Figure 1). This visual representation of knee, hip, and ankle joint angles in the sagittal plane highlights the interaction between surgery type and walking condition (preferred vs. fast-paced), represented as waveform differences. From these waveforms, discrete measures were analyzed to further evaluate these differences.

**Figure 1.**
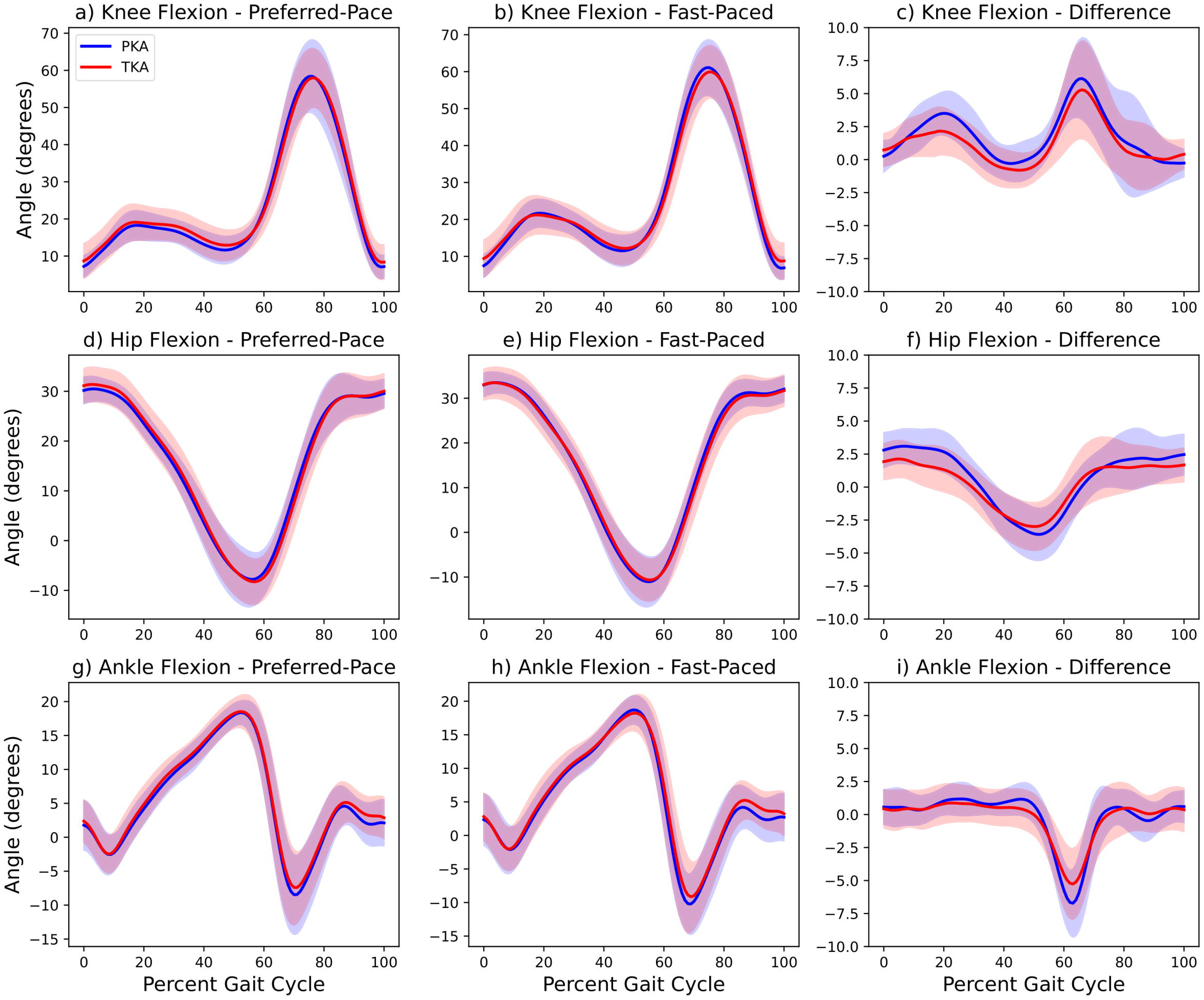
Ensemble waveforms with standard deviation shading for knee, hip, and ankle flexion angles during walking trials for PKA (blue) and TKA (red) patients on the surgical side. Sub-figures a), d), and g) show preferred-pace trials; b), e), and h) show fast-paced trials; and c), f), and i) show the computed difference between fast- and preferred-pace trials. Note: Difference waveforms (c, f, i) are plotted on a smaller y-axis scale to reflect the lower magnitude of change between conditions.

Linear mixed models were used to evaluate main effects of walk condition, and surgery type, along with the interaction of walk condition and surgery type for spatiotemporal parameters (Table 2). The main effect of walk condition had a statistically significant effect on gait speed (p < 0.001, effect size: 1.24) and stride length (p < 0.001, effect size: 0.98). Specifically, a change in walk condition from preferred to fast-pace increased gait speed and stride length for all patients, regardless of surgery type. However, the main effect of surgery type was not associated with spatiotemporal parameters overall. Additionally, the interaction of walk condition and surgery type revealed a statistically significant difference in stride length (p = 0.003, effect size: - 0.34). Specifically, the PKA group was found to have a greater increase in stride length when moving from preferred to fast-paced walking compared to the TKA group. Gait speed also increased more in the PKA group when moving from preferred to fast-paced walking compared to the TKA group; however, this did not reach statistical significance (p = 0.06, effect size: - 0.23).

**Table 2.**
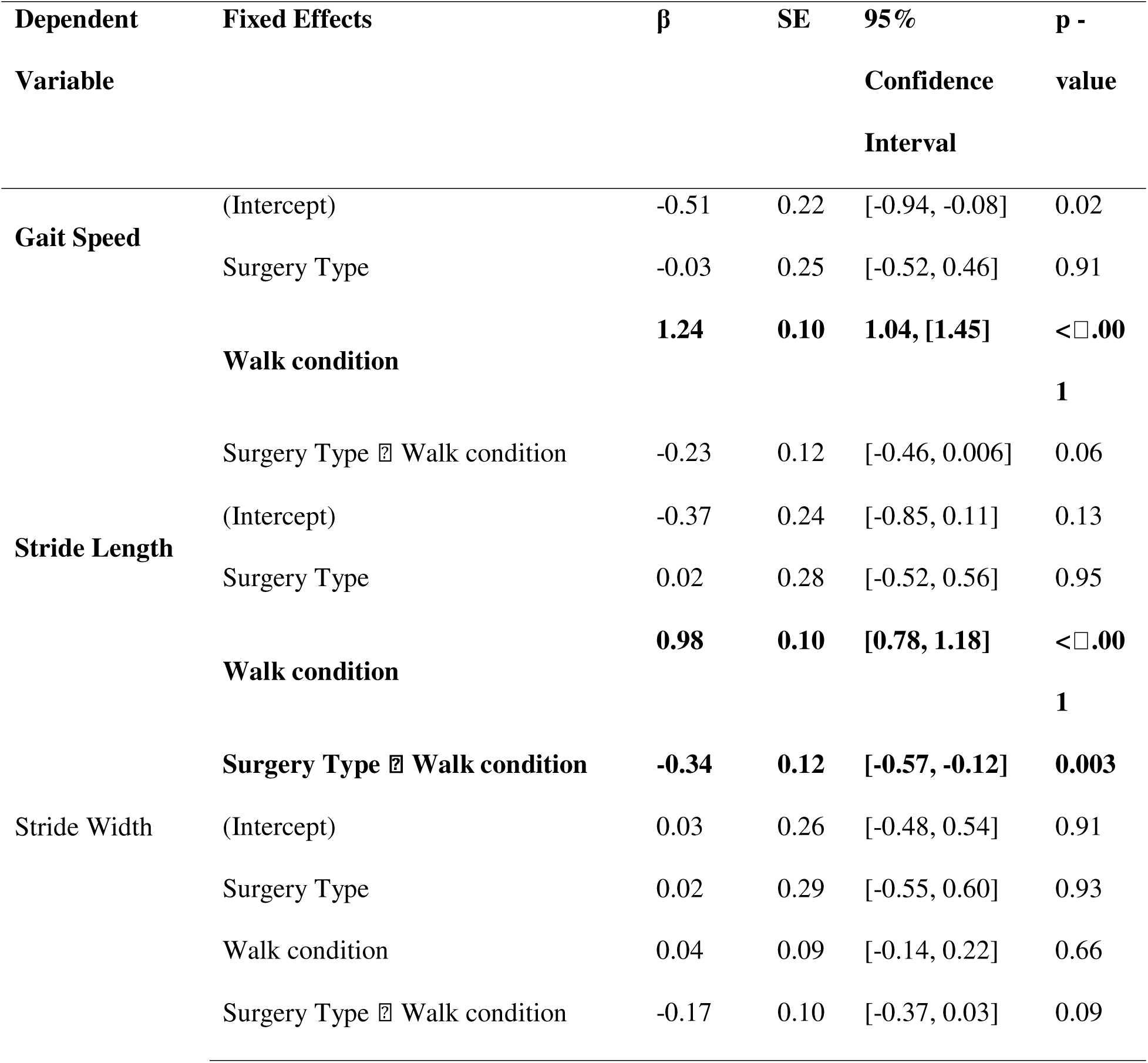
Linear Mixed Models for spatiotemporal parameters on surgical side. Statistically significant (p < 0.05) values in bold. Beta estimates are standardized; SE represents standard

| Dependent Variable | Fixed Effects | $\beta$ | SE | 95% Confidence Interval | p - value |
| --- | --- | --- | --- | --- | --- |
| Gait Speed | (Intercept) | -0.51 | 0.22 | [-0.94, -0.08] | 0.02 |
|  | Surgery Type | -0.03 | 0.25 | [-0.52, 0.46] | 0.91 |
|  | Walk condition | **1.24** | **0.10** | **1.04, [1.45]** | **<0.001** |
| | Surgery Type × Walk condition | -0.23 | 0.12 | [-0.46, 0.006] | 0.06 |
| Stride Length | (Intercept) | -0.37 | 0.24 | [-0.85, 0.11] | 0.13 |
|  | Surgery Type | 0.02 | 0.28 | [-0.52, 0.56] | 0.95 |
|  | Walk condition | **0.98** | **0.10** | **[0.78, 1.18]** | **<0.001** |
| | Surgery Type × Walk condition | **-0.34** | **0.12** | **[-0.57, -0.12]** | **0.003** |
| Stride Width | (Intercept) | 0.03 | 0.26 | [-0.48, 0.54] | 0.91 |
|  | Surgery Type | 0.02 | 0.29 | [-0.55, 0.60] | 0.93 |
|  | Walk condition | 0.04 | 0.09 | [-0.14, 0.22] | 0.66 |
| | Surgery Type × Walk condition | -0.17 | 0.10 | [-0.37, 0.03] | 0.09 |

Similar to spatiotemporal parameters, a main effect of walk condition exists for discrete gait measures of the knee (Table 3), hip (Table 4), and ankle (Table 5). Specifically, a change in walk condition from preferred to fast-pace increased peak stance knee flexion (p < 0.001, effect size: 0.71), peak swing knee flexion (p < 0.001, effect size: 0.75), knee excursion (p < 0.001, effect size: 0.85), peak stance hip flexion (p < 0.001, effect size: 0.87), peak swing hip flexion (p < 0.001, effect size: 0.64), hip ROM (p < 0.001, effect size: 1.02), and ankle ROM (p = 0.03, effect size: 0.30), and decreased peak stance hip extension (p < 0.001, effect size: -0.67), and peak ankle plantarflexion (p = 0.03, effect size: -0.26). However, the main effect of surgery type was not associated with discrete gait measures overall. Additionally, the interaction of walk condition and surgery type showed a statistically significant difference in all knee measures, some of the hip measures, and no ankle measures. The PKA group had a larger increase in peak knee flexion angle during stance phase (p = 0.004, effect size: -0.32), peak knee flexion angle during swing phase (p = 0.02, effect size: -0.50), and knee excursion (p <0 .001, effect size: -0.59) when moving from preferred to fast-paced walking compared to the TKA group on the surgical side. The PKA group also had a larger increase in peak hip flexion angle during stance phase (p = 0.02, effect size: -0.28), and hip ROM (p = 0.02, effect size: -0.29) when moving from preferred to fast-paced walking compared to the TKA group on the surgical side.

**Table 3.** Linear Mixed Models for knee discrete gait measures on surgical side. Statistically significant (p < 0.05) values in bold*. B*eta estimates are standardized; SE represents standard

| Dependent Variable | Fixed Effects | $\beta$ | SE | 95% Confidence Interval | p - value |
| --- | --- | --- | --- | --- | --- |
| **Knee Peak Stance Flexion** | (Intercept) | -0.35 | 0.25 | [-0.85, 0.15] | 0.17 |
|  | Surgery Type | 0.16 | 0.29 | [-0.41, 0.72] | 0.59 |
|  | **Walk condition** | **0.71** | **0.10** | **[0.52, 0.91]** | **<0.001** |
|  | **Surgery Type × Walk condition** | **-0.32** | **0.11** | **[-0.54, -0.11]** | **0.004** |
| **Knee Peak Swing Flexion** | (Intercept) | -0.15 | 0.26 | [-0.65, 0.35] | 0.55 |
|  | Surgery Type | -0.03 | 0.29 | [-0.59, 0.53] | 0.91 |
|  | **Walk condition** | **0.75** | **0.19** | **[0.38, 1.12]** | **<0.001** |
|  | **Surgery Type × Walk condition** | **-0.50** | **0.21** | **[-0.91, -0.08]** | **0.02** |
| **Knee Excursion** | (Intercept) | -0.07 | 0.25 | [-0.56, 0.43] | 0.80 |
|  | Surgery Type | -0.16 | 0.28 | [-0.72, 0.39] | 0.56 |
|  | **Walk condition** | **0.85** | **0.12** | **[0.62, 1.09]** | **<0.001** |
|  | **Surgery Type × Walk condition** | **-0.59** | **0.13** | **[-0.85, -0.33]** | **<0.001** |

**Table 4.**
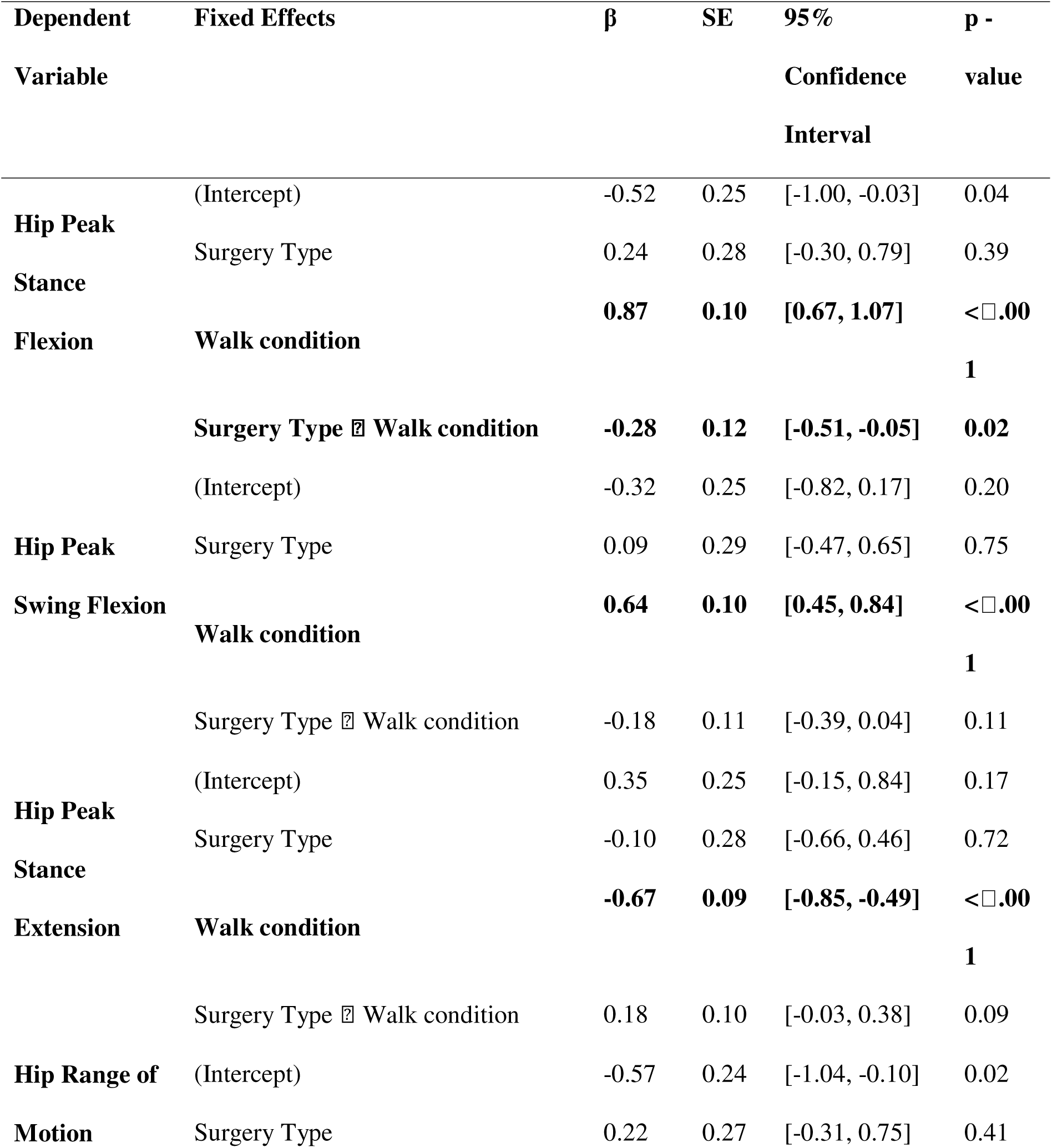
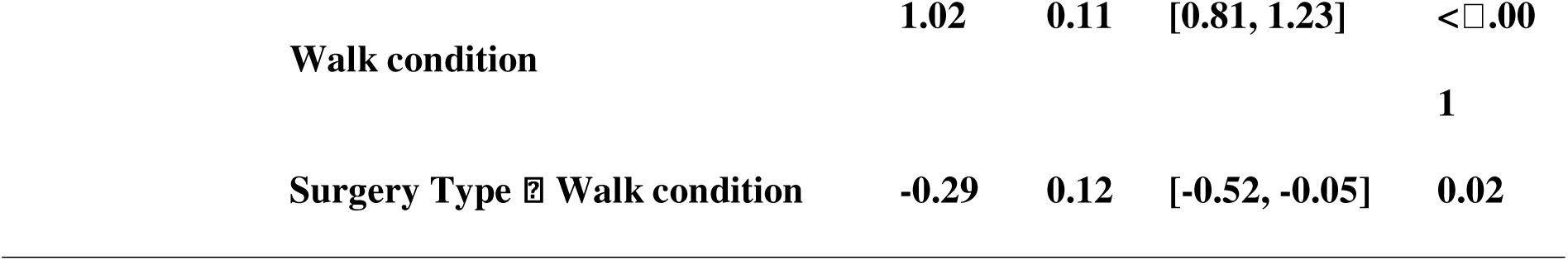
Linear Mixed Models for hip discrete gait measures on surgical side. Statistically significant (p < 0.05) values in bold. *B*eta estimates are standardized; SE represents standard

**Table 5.** Linear Mixed Models for ankle discrete gait measures on surgical side. Statistically significant (p < 0.05) values in bold*. B*eta estimates are standardized; SE represents standard

| **Dependent Variable** | **Fixed Effects** | **β** | **SE** | **95% Confidence Interval** | **p - value** |
| --- | --- | --- | --- | --- | --- |
| Ankle Peak | (Intercept) | -0.04 | 0.26 | [-0.56, 0.47] | 0.87 |
| Dorsiflexion | Surgery Type | 0.12 | 0.29 | [-0.46, 0.69] | 0.69 |
|  | Walk condition | 0.07 | 0.15 | [-0.23, 0.37] | 0.64 |
|  | Surgery Type × Walk condition | -0.21 | 0.17 | [-0.54, 0.13] | 0.22 |
| **Ankle Peak** | (Intercept) | 0.04 | 0.26 | [-0.47, 0.54] | 0.89 |
| **Plantarflexion** | Surgery Type | 0.15 | 0.29 | [-0.42, 0.72] | 0.61 |
|  | **Walk condition** | **-0.26** | **0.12** | **[-0.50, -0.03]** | **0.03** |
|  | Surgery Type × Walk condition | -0.06 | 0.14 | [-0.32, 0.21] | 0.68 |
| **Ankle Range of Motion** | (Intercept) | -0.06 | 0.26 | [-0.57, 0.45] | 0.82 |
|  | Surgery Type | -0.09 | 0.29 | [-0.66, 0.49] | 0.77 |
|  | **Walk condition** | **0.30** | **0.14** | **[0.03, 0.57]** | **0.03** |
|  | Surgery Type × Walk condition | -0.06 | 0.16 | [-0.36, 0.25] | 0.73 |

Student’s Independent Samples T-Tests and Mann-Whitney U Independent Samples T-Test were used to evaluate statistical significance of sit-to-stand measures (Table 6). There was no statistically significant difference found between PKA and TKA cohorts for mean time to complete all five trials, peak knee flexion, minimum knee flexion, and peak trunk flexion. Additionally, all sit-to-stand measures had small effect size.

**Table 6.** Sit-to-stand metrics showed no significant differences between cohorts. Values are reported as mean (standard deviation). For mean time, Mann-Whitney U Independent Samples T-Test and rank biserial correlation were used. For the remaining variables, Student’s Independent Samples T-Tests and Cohen’s d were used.

|  | PKA | TKA | p-value | Cohen's d |
| --- | --- | --- | --- | --- |
| Mean Time (s) | 15.91 (5.48) | 14.83 (4.44) | 0.75 | 0.05 |
| Peak Knee Flexion (°) | 66.01 (4.57) | 65.30 (4.95) | 0.62 | 0.15 |
| Minimum Knee Flexion (°) | 5.96 (2.99) | 7.18 (3.13) | 0.18 | -0.39 |
| Peak Trunk Flexion (°) | -41.56 (12.23) | -43.38 (13.13) | 0.63 | 0.14 |

## Discussion

The purpose of this study was to better understand the preoperative objective biomechanical differences between PKA and TKA cohorts, in addition to self-reported measures. As expected, no differences were observed when evaluating differences between cohorts through self-reported measures, nor at single gait speed conditions or sit-to-stand. However, when evaluating the differences in walking patterns between preferred and fast pace, the PKA group exhibited a significant change in stride length, peak knee flexion angles, and peak hip flexion angles between preferred and fast-paced walking compared to the TKA group. Overall, these results suggest that patients who eventually receive a PKA have a higher functional capacity pre-operatively, as they show evidence of higher adaptability to increasing gait speed.

Despite increasing interest in gait biomechanics after knee arthroplasty, few studies have examined preoperative differences between PKA and TKA. A systematic review was conducted by Dong and colleagues in 2023 to compare spatiotemporal, kinematic, and kinetic gait characteristics between PKA and TKA during level walking^24^. Of 13 included studies, all studies included spatiotemporal parameters, 5 studies included kinematics, and only 1 study evaluated preoperative kinematics^24^. While these studies mainly focused on postoperative comparisons between PKA and TKA, it is interesting to note some of the spatiotemporal and kinematic parameters that were found to be statistically significant between groups were similar to the parameters of this study that were found to be statistically significant preoperatively. Gait speed, stride length, and peak stance knee flexion were reported to be significantly higher in the PKA groups post-operatively^24^. While no direct comparison can be made between the results of this study and those of Dong and colleagues, these parallels underscore the value of incorporating preoperative biomechanical assessments in order to not only to establish meaningful baselines, but also to better understand the trajectory of these functional metrics postoperatively.

The only study that included a preoperative time point for biomechanical analysis during level walking included in the systematic review conducted by Dong and colleagues was a study by Braito et al^16^. They used a multivariate analysis of variance (MANOVA) to report on the main effects of surgical type and time, as well as the interaction between main effects^16^. Similar to the results of our study, Braito and colleagues found that spatiotemporal parameters including gait speed, stride length, and step width were not influenced by surgical type^16^. Additionally, they found no statistically significant differences between PKA and TKA in sagittal knee, hip, or ankle kinematics, regardless of timepoint^16^. These results match those of this study, where we found no differences between PKA and TKA cohorts when strictly analyzing single-paced biomechanical metrics. The waveforms in Figure 1 highlight similar patterns that agree with these results. When single-speed gait kinematics are observed, PKA and TKA candidates show similar sagittal knee, hip, and ankle kinematics. However, this study adds to Braito et al by evaluating multi-speed gait to look deeper into the possible differences between cohorts. We found that the PKA group increased stride length, along with many knee and hip kinematic parameters when comparing preferred and fast-paced walking. This is also evident in the waveforms of Figure 1, where plotting the differences between preferred and faster pace walking reveals differences between cohorts. Similar spatiotemporal and kinematic parameters have been reported to increase in healthy older adults in studies that evaluate multi-speed gait, including increased stride length, hip flexion during stance, and hip flexion during swing^29^. This further supports the suggestion that the PKA group may have higher functional capacity over the TKA group preoperatively, as the PKA group shows similar adaptability to healthy older adults when asked to increase their gait speed.

The results of this study provide a broader methodological framework for studying clinical gait populations. A critical takeaway from our findings is that meaningful biomechanical differences become apparent only when gait is assessed at multiple speeds, particularly when examining the changes between these speeds. These findings are consistent with previous work showing gait alterations in patients with moderate to severe knee OA compared to healthy controls^30^. Zeni and colleagues reported that, when walking speed was experimentally controlled to a single speed, the only significant biomechanical difference between OA patients and healthy participants was knee excursion^30^. However, additional differences emerged under fast walking conditions, including peak knee flexion, hip flexion, and ankle dorsiflexion^30^. This suggests that the modulation of gait speed is critical for revealing underlying functional impairments that may be masked under controlled or comfortable walking speeds. Our findings further suggest that evaluating the changes in biomechanical metrics between gait speeds provide valuable insight into functional deficits in OA populations.

In addition to multi-speed gait, our study investigated the kinematic differences in sit-to-stand tasks as this was expected to provide further insight into functional differences between cohorts that had not previously been studied^31^. A systematic review was conducted by Wang and colleagues to evaluate sit-to-stand outcomes following knee arthroplasty^31^. Of the 13 studies included, all involved either PKA or TKA; however, among the two studies including both cohorts, none directly compared PKA versus TKA outcomes^31^. Our results suggest that sit-to-stand performance may not meaningfully differentiate PKA and TKA patients.

There are some limitations to this study. Firstly, we evaluated discrete biomechanical measures as opposed to full waveform analysis. While our study provides results regarding maximum and minimum kinematic parameters, full waveform analysis such as a principal component analysis would evaluate characteristics of the full waveform. This method, however, is more challenging to interpret, and we therefore elected to perform a simpler analysis to build a foundational understanding of the dataset before pursuing more complex waveform approaches. Additionally, our study included an unequal sample size for PKA and TKA cohorts, with several more patients being in the TKA cohort. This makes it difficult to detect statistical differences between groups, since the smaller PKA group contributes less information and reduces the ability to identify true differences. However, this is a representative sample from the fracture clinic in which the study was conducted as notably less patients receive PKA. Finally, this study focuses only on preoperative data. Since this study did not evaluate post-operative conditions, we did not report on important surgical parameters such as assistive technology used, alignment techniques, or prosthetic details. Future studies should incorporate longitudinal data in order to identify how preoperative biomechanical data influences postoperative evaluation, as well as how these surgical parameters may influence outcomes.

## Conclusion

This study evaluated objective biomechanical differences between PKA and TKA cohorts, in addition to self-reported measures. These findings suggest that self-reported measures alone may not provide adequate insight into the functional differences between PKA and TKA groups preoperatively. Overall, these findings highlight that multi-speed gait assessments rather than single-speed walking or sit-to-stand tasks are likely a more sensitive approach for detecting biomechanical differences in OA patients and may have broader applicability to other clinical populations. Future research should integrate these objective biomechanical metrics, utilizing both preferred and fast-paced walking, to better understand changes in objective functional capacity perioperatively between PKA and TKA interventions.

## Data Availability

All data produced in the present study are available upon reasonable request to the authors.

## References

1. Statistics Canada. Prevalence of chronic diseases and risk factors among Canadians aged 65 years and older. Ottawa: Government of Canada; 2020.

2. Dillon CF, Rasch EK, Gu Q, Hirsch R. Prevalence of knee osteoarthritis in the United States: arthritis data from the Third National Health and Nutrition Examination Survey 1991–94. J Rheumatol 2006;33:2271–9.

3. Astephen JL, Deluzio KJ, Caldwell GE, et al. Biomechanical changes at the hip, knee, and ankle joints during gait are associated with knee osteoarthritis severity. J Orthop Res 2008;26:332–41.

4. Al-Zahrani KS, Bakheit AM. A study of the gait characteristics of patients with chronic osteoarthritis of the knee. Disabil Rehabil 2002;24:275–80.

5. Canadian Institute for Health Information. Hip and knee replacements in Canada: Canadian Joint Replacement Registry 2015 annual report. Ottawa: CIHI; 2020.

6. Kurtz S, Ong K, Lau E, et al. Projections of primary and revision hip and knee arthroplasty in the United States from 2005 to 2030. J Bone Joint Surg Am 2007;89:780–5.

7. Hatfield GL, Hubley-Kozey CL, Astephen Wilson JL, Dunbar MJ. The effect of total knee arthroplasty on knee joint kinematics and kinetics during gait. Clin Orthop Relat Res 2011;26:309–18.

8. Marino G, De Capitani F, Adamo P, et al. Long-term gait analysis in patients after total knee arthroplasty: a systematic review and meta-analysis. Gait Posture 2024;113:75–98.

9. Stoddart JC, Dandridge O, Garner A, et al. The compartmental distribution of knee osteoarthritis: a systematic review and meta-analysis. Osteoarthr Cartil 2021;29:445–55.

10. Beard DJ, Davies LJ, Cook JA, et al. Total versus partial knee replacement in patients with medial compartment knee osteoarthritis: the TOPKAT RCT. Health Technol Assess 2020;24:1–98.

11. Chawla JP, van der List JP, Christ AB, et al. Annual revision rates of partial versus total knee arthroplasty: a comparative meta-analysis. Knee 2017;24:179–90.

12. Smith S, Alvand A, Locock L, et al. Partial or total knee replacement? Identifying patients’ information needs on knee replacement surgery: a qualitative study to inform a decision aid. Qual Life Res 2020;29:999–1011.

13. Liddle AD, Pandit H, Judge A, Murray DW. Patient-reported outcomes after total and unicompartmental knee arthroplasty. Bone Joint J 2015;97-B:793–801.

14. Amin AK, Patton JT, Cook RE, et al. Unicompartmental or total knee arthroplasty? Results from a matched study. Clin Orthop Relat Res 2006;451:101–6.

15. Agarwal A, Miller S, Hadden W, et al. Comparison of gait kinematics in total and unicondylar knee replacement surgery. Ann R Coll Surg Engl 2019;101:391–8.

16. Braito M, Giesinger JM, Fischler S, et al. Knee extensor strength and gait characteristics after minimally invasive unicondylar knee arthroplasty vs minimally invasive total knee arthroplasty: a nonrandomized controlled trial. J Arthroplasty 2016;31:1711–6.

17. De Vroey H, Staes F, Vereecke E, et al. Lower extremity gait kinematics outcomes after knee replacement demonstrate arthroplasty-specific differences between unicondylar and total knee arthroplasty: a pilot study. Gait Posture 2019;73:299–304.

18. Çankaya D. Unicompartmental knee arthroplasty results in a better gait pattern than total knee arthroplasty: gait analysis with a smartphone application. Jt Dis Relat Surg 2021;32:22–7.

19. Copp EH, Gale TH, Byrapogu VKC, et al. Unicompartmental knee arthroplasty approximates healthy knee kinematics more closely than total knee arthroplasty. J Orthop Res 2024;42:2514–24.

20. Friesenbichler B, Item-Glatthorn JF, Wellauer V, et al. Short-term functional advantages after medial unicompartmental versus total knee arthroplasty. Knee 2018;25:638–43.

21. Pongcharoen B, Liengwattanakol P, Boontanapibul K. Comparison of functional recovery between unicompartmental and total knee arthroplasty: a randomized controlled trial. J Bone Joint Surg Am 2023;105:191–201.

22. Garner AJ, Dandridge OW, Richard R, et al. The compartmental approach to revision of partial knee arthroplasty results in nearer-normal gait and improved patient reported outcomes compared to total knee arthroplasty. Knee Surg Sports Traumatol Arthrosc 2021;31:1143–52.

23. Sershon RA, Fricka KB, Hamilton WG, et al. Early results of a randomized controlled trial of partial versus total knee arthroplasty. J Arthroplasty 2022;37:S94–S97.

24. Dong M, Fan H, Yang D, et al. Comparison of spatiotemporal, kinematic, and kinetic gait characteristics in total and unicompartmental knee arthroplasty during level walking: a systematic review and meta-analysis. Gait Posture 2023;104:58–69.

25. Dawson J, Fitzpatrick R, Murray D, Carr A. Questionnaire on the perceptions of patients about total knee replacement. J Bone Joint Surg Br 1998;80:63–9.

26. EuroQol Research Foundation. EQ-5D-5L User Guide. Rotterdam: EuroQol Research Foundation; 2019. Available from: https://euroqol.org/publications/user-guides

27. Kroenke K, Strine TW, Spitzer RL, Williams JB, Berry JT, Mokdad AH. The PHQ-8 as a measure of current depression in the general population. J Affect Disord 2009;114:163–73.

28. Outerleys J, Mihic A, Keller V, Laende E, Deluzio K. Markerless motion capture provides repeatable gait outcomes in patients with knee osteoarthritis. J Biomech 2024;168:112115.

29. Fukuchi CA, Fukuchi RK, Duarte M. Effects of walking speed on gait biomechanics in healthy participants: a systematic review and meta-analysis. Syst Rev 2019;8:153.

30. Zeni JA Jr, Higginson JS. Differences in gait parameters between healthy subjects and persons with moderate and severe knee osteoarthritis: a result of altered walking speed? Clin Biomech (Bristol, Avon) 2009;24:372–8.

31. Wang J, Siddicky SF, Oliver TE, Dohm MP, Barnes CL, Mannen EM. Biomechanical changes following knee arthroplasty during sit-to-stand transfers: systematic review. J Arthroplasty 2019;34:2494–2501.

